# PI-RADS v2 is a Strong Prognostic Marker for Adverse Outcomes in Prostate Cancer

**DOI:** 10.1101/2025.03.20.25324337

**Authors:** Tolou Shadbahr, Juho Pylväläinen, Juho Eineluoto, Alessio Moro, Oleg Kerro, Anu Kenttämies, Eugen Czeizler, Teemu J Murtola, Hyon-Jung Kim-Ollila, Philippe Puech, Jonathan Olivier, Arnauld Villers, Jaakko Peltonen, Matti Nykter, Jing Tang, Tuomas Mirtti, Teemu D. Laajala, Antti S. Rannikko

## Abstract

**Importance:** Magnetic Resonance Imaging (MRI) coupled with Prostate Imaging-Reporting and Data System (PI-RADS) provides a standardized scoring system for assessing the clinical significance of prostate cancer (PCa). The association between PI-RADS scores and key clinical end-points remains underexplored due to limited follow-up data.

**Objective:** To evaluate the association of PI-RADS with prostate cancer-specific mortality (PCSM), overall survival (OS), metastasis-free survival (MFS), and biochemical recurrence (BCR) across three cohorts.

**Design, Setting, Participants:** A retrospective registry cohort, Helsinki University Hospital (HUS) for men with clinical suspicion of PCa, included 4,674 men who underwent diagnostic MRI during the PI-RADS version-2 era (2015-2019, median follow-up [mFU] 5.0 years). Validation cohorts were: Tampere University Hospital (n=1,474, 2016-2021, mFU 2.2 years) and Lille University Hospital (N=301, 2016-2024, mFU 6.2 years).

**Exposures:** Diagnostic MRI imaging was scored using PI-RADS v2.

**Main outcomes and measures:** Primary outcomes were PCSM, OS, MFS, and BCR. Secondary measures included clinical confounders, such as age, Grade Group (GG), and Charlson’s Comorbidity Index (CCI).

**Results:** 84 of 4,674 HUS-cohort patients (1.7%) died of PCa, with 79 patients (94%) presenting with PI-RADS score 5. Multivariable Cox regression adjusted for clinical confounders found PI-RADS score 5 was significantly associated with PCSM (Hazard Ratio (HR) 18.4, 95% CI [6.62-51.1], p<0.001), along with biopsy GG 5 (HR 5.45 [1.82-16.3], p=0.002), CCI (HR 1.53 [1.42-1.7], p<0.001), and log2-PSA (HR 1.28 [1.11-1.5], p<0.001). Kaplan-Meier curves demonstrated negligible PCSM after negative MRI (nMRI; PI-RADS score 0-2) (log-rank p<0.001). PI-RADS score 5 was associated with OS in the Helsinki and Tampere cohorts (p<0.001) and MFS and BCR in the Lille cohort (p<0.001).

**Conclusion and relevance:** PI-RADS score 5 is strongly associated with an increased risk of PCSM and other clinically relevant outcomes. A non-suspicious MRI associates with a negligible risk of adverse outcomes. These findings support the use of MRI for risk stratification in PCa.

## Introduction

Prostate cancer (PCa) is the second most frequently diagnosed malignancy in men and fifth cause of death in men globally.^1^ Prostate-specific antigen (PSA) testing and transrectal ultrasound (TRUS) guided biopsy are widely used diagnostic tools that have been shown to reduce prostate cancer-specific mortality (PCSM) in screening. However, these diagnostic pathways have also contributed to overdiagnosis and overtreatment.^2^ These drawbacks can be mitigated through the adoption of MRI-based diagnostics combined with the standardized Prostate Imaging Reporting and Data System (PI-RADS).^3–12^

MRI-based diagnostic approaches (MRI-targeted biopsies and MRI-imaging) have demonstrated superior accuracy in detecting high-grade cancers compared to systematic biopsy, in screening ^13–15^ and in cases of clinical suspicion.^10,16^ Epidemiological ^17,18^, histopathological ^19,20^, and genetic ^21,22^ studies suggest that MRI-invisible lesions may be biologically less aggressive than MRI-visible lesions of the same grade. However, Long-term, real-world validation of PI-RADS scoring has been limited, as clinical adoption only began in 2012 for PI-RADS version 1^3^ and in 2015 version 2 (v2)^4^.

PI-RADS standardizes the acquisition of prostate MRI images, their interpretation, and reporting of their findings. Early studies have linked positive MRI findings (PI-RADS scores 3 to 5) with an increased risk of clinically significant PCa, a greater likelihood of positive surgical margins, and a higher risk of biochemical recurrence (BCR) after radical prostatectomy (RP).^6,19,23–29^ Recently, Wibmer et al. highlighted a discontinuity in relating MRI findings with the strongest clinical end-points, such as PCSM ^24,30^. To address the lack of PI-RADS era cohorts, Wibmer and colleagues retrospectively assigned PI-RADS scores to the pre-PI-RADS era MRIs and concluded that MRI findings are associated with long-term clinicopathologic outcomes. However, these retrospective scores were based on anatomical sequences, excluding key PI-RADS functional sequences such as Diffusion-Weighted Imaging, which are crucial for contemporary MRI in clinical practice.

This multicohort study evaluates the association of PI-RADS v2 with key clinical outcomes in PCa, including PCSM, overall survival (OS), metastasis-free survival (MFS), and BCR.

## Materials and Methods

All study patients underwent multiparametric MRI (mpMRI) during the PI-RADS v2 era. Table 1 summarizes all cohorts and their key variables.

**Table 1.** Patient demographics for the three cohorts. Count data is shown as patient amounts (percentage inside column) and numeric data as median (IQR).

| Cohort |  | Helsinki | Tampere | Lille |
| --- | --- | --- | --- | --- |
| N |  | 4674 | 1474 | 301 |
| Age at MRI (years) |  | 68 (61 - 73) | 67 (62 - 72) | 68 (64 - 84) |
| Follow-up (years) |  | 5.0 (4.2 - 5.8) | 2.21 (1.11 - 3.44) | 6.2 (4.8 - 7.4) |
| Pre-MRI PSA |  | 7.8 (5.1 - 11.7) | 7.0 (5.0 - 11.0) | 6.9 (5.3 - 10.0) |
| Negative MRI | PI-RADS score 0 | 1555 (33%) | - | - |
|  | PI-RADS score 1 | 7 (0%) | 81 (6%) | - |
|  | PI-RADS score 2 | 51 (1%) | 191 (13%) | 14 (5%) |
| Positive MRI | PI-RADS score 3 | 718 (15%) | 224 (15%) | 43 (14%) |
|  | PI-RADS score 4 | 928 (20%) | 607 (41%) | 96 (32%) |
|  | PI-RADS score 5 | 1415 (30%) | 371 (25%) | 148 (49%) |
| Biopsy | Benign | 831 (18%) | - | - |
|  | GG 1 | 375 (8%) | 511 (35%) | 62 (21%) |
|  | GG 2/3 <sup>a</sup> | - | 23 (2%) | - |
|  | GG 2 | 726 (16%) | 400 (27%) | 132 (44%) |
|  | GG 3 | 519 (11%) | 154 (10%) | 86 (29%) |
|  | GG 4 | 214 (5%) | 97 (7%) | 15 (5%) |
|  | GG 5 | 311 (7%) | 166 (11%) | 6 (2%) |
|  | No biopsy or unknown | 1698 (36%) | 123 (8%) | - |
| CCI | 0 | 3638 (78%) | - | - |
|  | 1-3 | 871 (19%) | - | - |
|  | 4+ | 160 (3%) | - | - |
| Interventions | No known interventions or only non-curative treatments | 2671 (57%) | 526 (36%) | - |
|  | RP | 706 (15%) | 324 (22%) | 301 (100%) |
|  | RT | 1053 (23%) | 573 (39%) | - |
|  | RP + RT | 244 (5%) | 51 (3%) | - |
| Event type | Censoring | 4216 (90%) | 1444 (98%) | 275 (91%) |
|  | Non-PCa death | 374 (8%) | - | 24 (8%) |
|  | PCa death | 84 (2%) | - | 2 (1%) |
|  | Any death (PCa or Non-PCa) | - | 30 (2%) | - |

CCI: CCI: (adjusted) Charlson's Comorbidity Index
GG: ISUP Grade Group
RT: Radiation Therapy
RP: Radical Prostatectomy
<sup>a</sup>: For some registry data, only Gleason sum 7 information was available

### Helsinki University Hospital Study Cohort

Helsinki-Uusimaa Hospital District encompasses the largest tertiary care patient population in Finland. Registries from Helsinki University Hospital (HUS) were utilized to integrate MRI imaging data, clinical variables, and adverse outcomes. The HUS cohort included men undergoing diagnostic evaluation for suspected PCa (Figure 1A). Patient exclusion criteria are outlined in Supplementary Figure S1.

**Figure 1.**
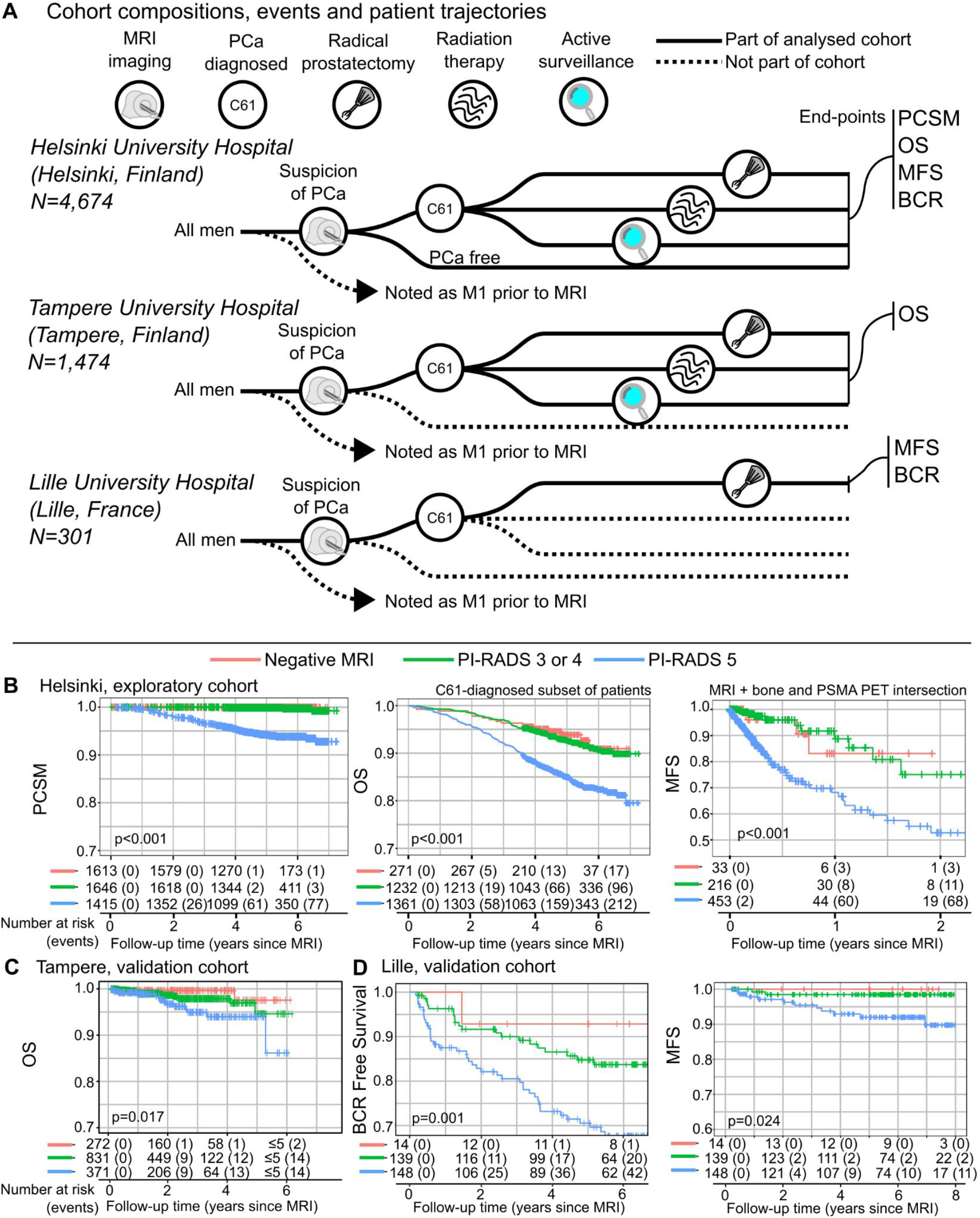
(A) Schematic overview of patient cohort composition in Helsinki, Tampere, and Lille; Kaplan-Meier curves and numbers at risk with events for (B) Helsinki cohort PCSM, OS for patients with PCa diagnosis, and MFS; (C) Tampere cohort OS for in-hospital death; and (D) Lille cohort BCR / Relapse Free Survival and MFS. Reported p-values are log-rank tests.

A total of 4,674 patients were included, with a median follow-up time of 5.0 years (IQR 4.2-5.8). Follow-up began on the date of the MRI recorded in the HUS electronic health record (EHR) system (HUS Acamedic). mpMRIs were performed at HUS between 2015 and 2019, following the adoption of the PI-RADS v2, with imaging protocols described in Pylväläinen et al.^18^ Radiologists with over five years of experience in prostate MRI interpreted the images according to the European Society of Urogenital Radiology PI-RADS v2 guidelines.

An additional pre-PI-RADS era MRI cohort from HUS was identified to allow for longer follow-up despite the lack of PI-RADS reporting (Supplementary Methods).

Patients with metastatic disease at baseline or with biopsy results recorded outside a one-year window from the first MRI were excluded (Supplementary Figure S1). The ISUP Grade Group (GG) of biopsies was extracted from semi-structured and free-form pathological statements. Among patients with unknown biopsy status, the majority were confirmed to have nMRI findings (Supplementary Table S1).

Table 2 provides a comprehensive overview of HUS cohort variables stratified by PI-RADS scores. Primary outcomes included PI-RADS scores recorded during the v2 era and clinical end-points such as PCSM, OS, MFS, and BCR. Cause of death was retrieved from the national Statistics Finland registry, which has excellent agreement (kappa 0.95), with 96% sensitivity and 99% specificity for identifying prostate cancer deaths.^31^ Secondary measurements included clinical confounders relevant to PCa, such as PSA levels, patient age, biopsy GGs, known interventions, and comorbidities (Table 1).

**Table 2.** Patient demographics in PI-RADS version 2 era in HUS, with stratification to negative MRI findings (PI-RADS 0 to 2), PI-RADS score 3 to 4, and PI-RADS score 5. Count data is shown as patient amounts (percentage inside column) and numeric data as median (IQR).

|  |  | All PI-RADS | Negative MRI | PI-RADS 3 & 4 | PI-RADS 5 |
| --- | --- | --- | --- | --- | --- |
| **N** |  | 4674 | 1613 | 1646 | 1415 |
| **Age at MRI (years)** |  | 68 (61 - 73) | 65 (59 - 71) | 68 (62 - 73) | 70 (64 - 75) |
| **Follow-up (years)** |  | 5.0 (4.2 - 5.8) | 4.8 (4.1 - 5.6) | 5.1 (4.2 - 6.0) | 5.0 (4.1 - 6.0) |
| **Pre-MRI PSA (µg/L)** |  | 7.8 (5.1 - 11.7) | 6.6 (4.0 - 9.7) | 7.3 (5.0 - 10.1) | 10.3 (7.2 - 17.6) |
| **Biopsy** | **Benign** | 831 (18%) | 349 (22%) | 399 (24%) | 83 (6%) |
|  | **GG 1** | 375 (8%) | 53 (3%) | 231 (14%) | 91 (6%) |
|  | **GG 2** | 726 (16%) | 53 (3%) | 362 (22%) | 311 (22%) |
|  | **GG 3** | 519 (11%) | 19 (1%) | 190 (12%) | 310 (22%) |
|  | **GG 4** | 214 (5%) | 5 (0%) | 71 (4%) | 138 (10%) |
|  | **GG 5** | 311 (7%) | 6 (0%) | 64 (4%) | 241 (17%) |
|  | **No biopsy or unknown** | 1698 (36%) | 1128 (70%) | 329 (20%) | 241 (17%) |
| **CCI** | **0** | 3922 (77%) | 1262 (78%) | 1320 (80%) | 1056 (75%) |
|  | **1-3** | 981 (19%) | 312 (19%) | 291 (18%) | 268 (19%) |
|  | **4+** | 205 (4%) | 35 (2%) | 34 (2%) | 91 (6%) |
| **Interventions** | **No known interventions or only non-curative treatments** | 2671 (57%) | 1509 (94%) | 890 (54%) | 272 (19%) |
|  | **RP** | 706 (15%) | 56 (3%) | 327 (20%) | 323 (23%) |
|  | **RT** | 1053 (23%) | 41 (3%) | 343 (21%) | 669 (47%) |
|  | **RP + RT** | 244 (5%) | 7 (0%) | 86 (5%) | 151 (11%) |
| **Event type** | **Censoring** | 4216 (90%) | 1519 (94%) | 1513 (92%) | 1184 (84%) |
|  | **Non-PCa death** | 374 (8%) | 93 (6%) | 129 (8%) | 152 (11%) |
|  | **PCa death** | 84 (2%) | 1 (0%) | 4 (0%) | 79 (6%) |

CCI: (adjusted) Charlson's Comorbidity Index
GG: ISUP Grade Group
RT: Radiation Therapy
RP: Radical Prostatectomy

PSA levels were obtained from HUS-laboratory services (HUSLAB). For patients with multiple PSA measurements, the most recent value at the time of the MRI was selected. PSA values were missing for 779 patients and were imputed using multiple imputation by chained equations (MICE). Quan’s adjusted Charlson’s Comorbidity Index (CCI) was calculated using all ICD-10 diagnosis codes available in the EHR database up to the time of MRI, with an additional three-month window. PCa diagnosis code (C61) was excluded from the CCI calculations to assess the comorbidity burden independent of confirmed PCa diagnosis.

Metastasis status was not categorically reported but was collected for patients who underwent bone scans or PSMA PET scans at HUS. Data were crosslinked to two separate ongoing studies. Metastasis screening was performed at the clinician’s discretion, typically in men with higher risk disease. Metastatic disease (M1) was defined as at least one positive lesion in the lymph node, soft tissue or bone in the Lille and HUS cohorts. Among 864 patients of the HUS cohort who underwent bone or PET scans due to suspected metastases, 741 were confirmed as metastasis-free (N0M0), and 123 patients were identified as having metastatic disease. Further details regarding metastasis data collection are provided in the supplementary material.

The use of registry data for this study was approved by the HUS Institutional Board (HUS/333/2019). The research adhered to the World Medical Association’s Declaration of Helsinki on good research practices. As this study was based on registry data, no explicit patient consent was required.

### Tampere University Hospital and Lille University Hospital Validation Cohorts

Pirkanmaa Hospital District (Tampere University Hospital) is the second-largest hospital district in Finland. Between 2000 and 2021, 17,125 men were diagnosed with PCa (ICD-10 code C61). Their PI-RADS scores, PSA values, and biopsy GGs were retrospectively collected from Tampere University Hospital EHR system (Figure 1A). Patients were excluded when they had missing birthdate, or missing follow-up information (e.g., any-cause death) and follow-up of less than 28 days post-MRI. After applying these criteria, 1,474 patients were included in the Tampere cohort, with a median follow-up time of 2.2 years (IQR 1.1-3.4). The recorded endpoint was any-cause mortality.

The Lille University Hospital cohort originally included 713 men who underwent MRI between 2007 and 2022. A subset of 301 patients with PI-RADS v2 MRI reports was selected, restricted to those who underwent MRI imaging after 2015 (Figure 1A). The median follow-up time was 6.2 years (IQR 4.8-7.4). All patients in the Lille cohort underwent RP, and the primary endpoints included metastasis-free survival and biochemical recurrence-free survival. For more details on the Lille cohort and its selection criteria, refer to Bommelaere et al.^32^. Additional details of the Tampere and Lille cohorts are provided in the Supplementary Material.

### Statistical analyses

Statistical analyses, visualizations, and data processing were conducted using R Statistical Software (R Core Team, 2024; Vienna, Austria; version 4.4.0 and newer) and Python (Python Software Foundation; version 3.8.0 and newer). Imputation was performed using the MICE R-package (version 3.16.0)^33^ and CCI was calculated using comorbidity R-package (v1.1.0). Kaplan-Meier curves were generated using R-packages survival (v3.8.3), ggsurvfit (v1.1.0), and survminer (v0.5.0). Competing risks were visualized using CompSurv (v0.1.0) via cumulative proportions over time, categorized by patient statuses alive, non-PCa death, PCa-specific death, and censoring (lost to follow-up). These visualizations were inspired by Albertsen et al.^34^ but included censoring to account for incomplete event follow-up data. Fine-Grey competing risk models were fit using tidycmprsk R-package (v1.1.0). Multivariable Cox proportional hazards models were used to evaluate the association between PI-RADS scores and disease endpoints, adjusting for clinical confounders including patient’s age at MRI, PSA, biopsy GG, interventions, and CCI. Hazard Ratios (HRs), 95% Confidence Intervals (CIs), and p-values were reported, with p<0.05 considered statistically significant in all analyses.

## Results

### Association Between PI-RADS Scores and Clinically Relevant Outcomes

We first evaluated the association between PI-RADS scores and clinically relevant outcomes Kaplan-Meier analysis of the HUS cohort revealed that men with a PI-RADS score of 5 had an elevated risk of PCSM, all-cause mortality, and metastasis (Figure 1B; p<0.001). After a median follow-up of 2.8 years, 3% of men with PI-RADS 5 experienced PCSM. A similar statistically significant trend was observed for PI-RADS 5 in association with OS and MFS.

When PI-RADS 5 was combined with GG 5, representing very high-risk aggressive prostate cancer, PCSM increased to 13% (32/241) within four years (Supplementary Figure S2; p<0.001).

In the Tampere validation cohort, PI-RADS 5 was significantly associated with OS trends (Figure 1C; p<0.05). In the Lille validation cohort of RP-treated men, PI-RADS 5 was significantly associated with MFS and BCR (Figure 1D; p<0.005). An additional pre-PI-RADS era cohort from Helsinki, with a median follow-up of 10.2 years, demonstrated a significant difference in PCSM risk between negative versus positive MRI findings (Supplementary Figure S3; p<0.001).

### Competing risk analysis

We explored competing risks during follow-up by plotting patient proportions over time, categorized as alive, censored, PCa-specific death and non-PCa death over a six-year-follow period in the HUS cohort (Figure 2). PI-RADS and biopsy GG were both associated with PCSM. Men with PI-RADS 5 lesions had a significantly higher risk of PCSM compared to men with PI-RADS 0-4. PCa-specific deaths remained consistent across age groups, but age was associated with an increased non-PCa mortality, as expected (Supplementary Figure S4).

**Figure 2.**
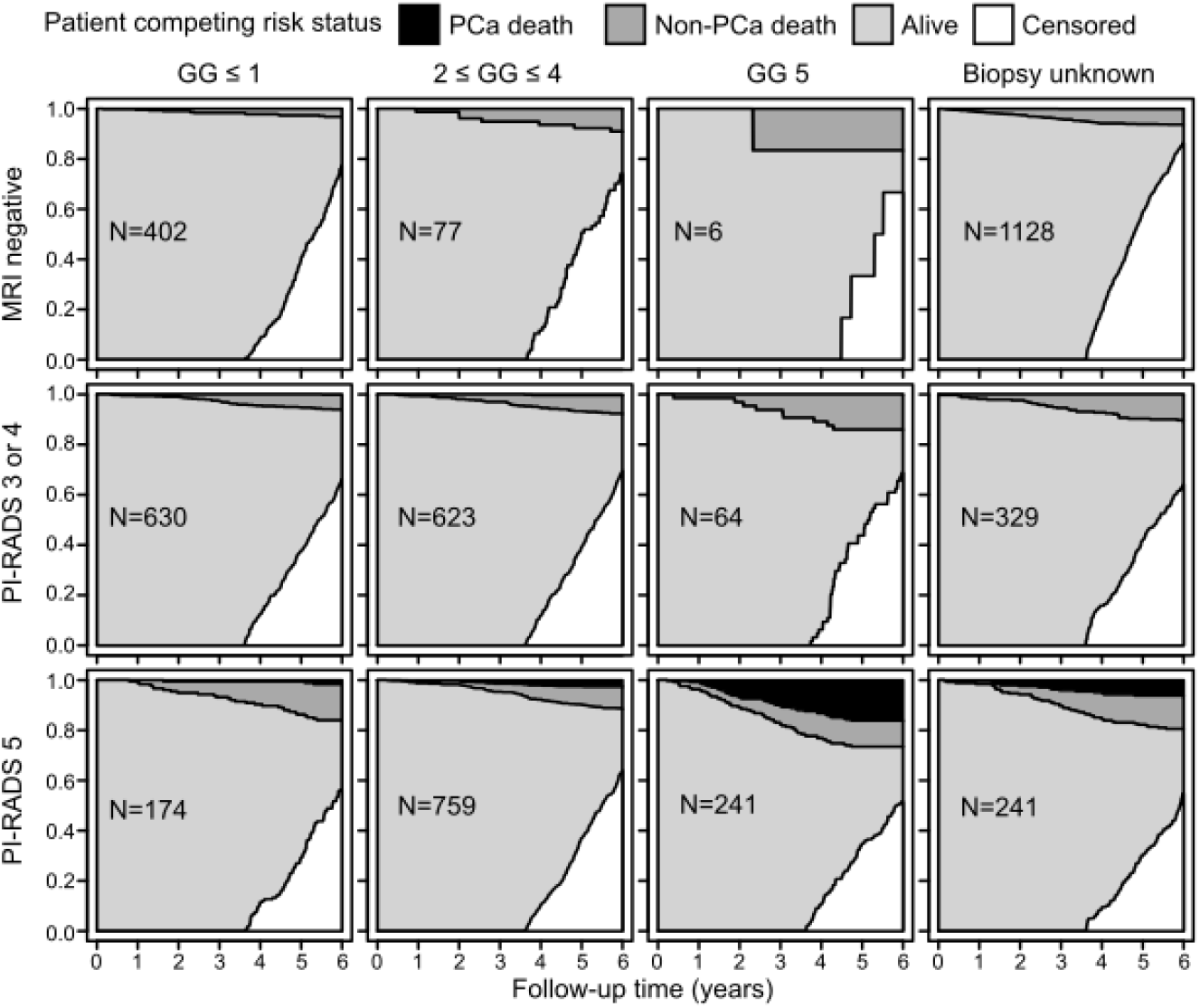
Patient proportions during follow-up for competing risks with alive, censored, non-PCa death and PCa-specific mortality shown as a function of time in HUS cohort.

We fitted PCa-specific death and non-PCa death into a Fine-Gray model thus treating them as competing endpoints for PI-RADS and biopsy GG (Supplementary Table S2). PI-RADS score 5 was highly associated with PCa-death (HR 75.1; CI 95% [10.2, 554], p<0.001) together with GG 5 (HR 10.1 [3.63, 28.1], p<0.001) and GG Unknown (HR 3.50 [1.19, 10.3], p=0.023). PI-RADS score 3 or 4 (HR 1.55 [1.15, 2.07], p=0.004) and PI-RADS 5 (HR 2.03 [1.49, 2.79], p<0.001) and GG Unknown (HR 1.60 [1.20, 2.13], p=0.002) were also associated with elevated risk of non-PCa-mortality, albeit with lower hazard ratios.

### Multivariable Analysis Adjusting for Clinical Confounders

We performed multivariable Cox regression analysis in the HUS cohort to adjust for confounders and to assess whether PI-RADS score 5 was still significantly associated with PCSM (Figure 3A) and OS (Figure 3B). For PCSM (Figure 3A), PI-RADS score 5 remained a strong prognostic indicator (HR 18.40; 95% CI [6.62-51.1], p<0.001), along with log2-transformed PSA (HR 1.28 [1.11-1.5], p<0.001), GG 5 (HR 5.45 [1.83-16.3], p=0.002), and CCI (HR 1.53 [1.42-1.7], p<0.001). Immediate curative-intent interventions (RP, RT) were not significant predictors of PCSM (HR 0.70 [0.36-1.4], p=0.284).

**Figure 3.**
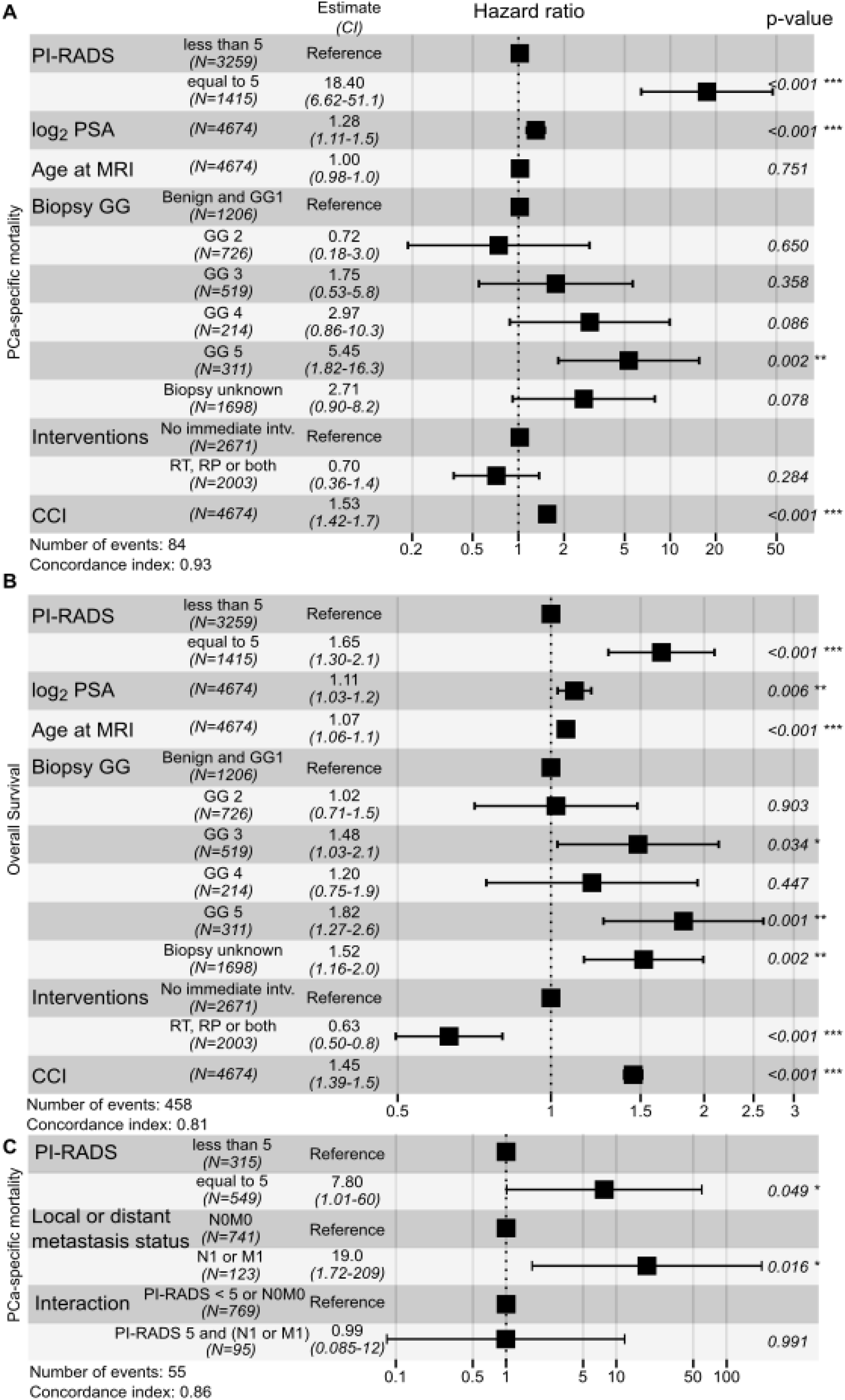
Multivariable Cox model forest plot in PI-RADS version 2 era with hazard ratios (95% CI) for (A) PCa-specific mortality; (B) Overall Survival; (C) PCa-specific mortality for combining PI-RADS and patients’ metastatic status at the time of diagnostic MRI.

For OS (Figure 3B), PI-RADS 5 (HR 1.65 [1.30-2.1], p<0.001), log2-transformed PSA (HR 1.11 [1.03-1.2], p=0.006), GG 5 (HR 1.82 [1.27-2.6], p<0.001), and CCI (HR 1.45 [1.39-1.5], p<0.001) were significantly associated with an increased risk of death. As expected, age at MRI was also significantly associated with increased all-cause mortality (HR 1.07 [1.06-1.1], p<0.001). Additionally, an unknown biopsy status was linked to higher all-cause mortality (HR 1.52 [1.16-2.0], p=0.002), which probably reflected patient selection bias.

### PI-RADS 5 and metastatic spread

We used Cox models for PCSM (Figure 3C) to explore the relationship between PI-RADS 5 and metastatic spread. Both PI-RADS and N/M-status were significant independent explanatory variables (p<0.05). However, their interaction was not significant. This trend remained consistent even when biopsy GG was incorporated into the multivariable Cox model (Supplementary Figure S5).

## Discussion

Our main finding is that a PI-RADS score 5 is associated with a significantly elevated risk of prostate cancer-specific mortality. This association remains robust after adjusting for confounders and extends consistently to other critical clinical endpoints, including OS, MFS, and BRC across multiple cohorts. Conversely, our data suggest that an nMRI is associated with a very low risk of adverse outcome.

To our knowledge, this is the first study to directly assess PI-RADS scores at the time of diagnosis in relation to PCSM using PI-RADS v2 era data now that prostate MRI has been adopted as a standard pre-biopsy triaging tool for men suspected of having localized or locally advanced non-metastatic PCa.

We specifically focused on PI-RADS v2, as it best represents current guideline recommendations for MRI as a pre-biopsy triage test. We acknowledge that this choice limits the follow-up duration, as PI-RADS and PI-RADS v2 recommendations were introduced in 2012 and 2015, respectively. To address this, we extended our analysis to a pre-PI-RADS era cohort, where MRI results were classified as either positive or negative, rather than using the five-tier PI-RADS system. Our findings were consistent: men with a positive MRI had a significantly higher risk of PCSM and other adverse clinical endpoints, whereas men with an nMRI had a negligible risk.

Our results align with the findings by Wibmer et al. who analyzed a cohort with T2-weighted MRI images in the pre-PI-RADS era and reported that MRI findings correlated with PCSM ^24,30^. Our data suggest that in addition to contemporary multiparametric-MRI or biparametric-MRI incorporating diffusion-weighted imaging and possible dynamic contrast-enhanced imaging; anatomical sequences alone may also contain some prognostic biomarkers. Future studies should further explore the prognostic value of prostate MRI, integrating both anatomical and functional imaging sequences to enhance risk stratification and deepen our understanding of PCa pathogenesis.

Interestingly, our findings indicate that prognosis may be primarily driven by MRI-defined tumor size and invasive behavior, rather than diffusion imaging signal intensity. According, PI-RADS guidelines^4^, tumor size (the greatest dimension) and invasive behavior are what distinguish PI-RADS 4 from PI-RADS 5. Further studies are needed to confirm this, and it is of interest whether radiomics models can add prognostic value beyond the PI-RADS classification.

We validated our findings with two independent external cohorts. In Tampere, Finland, we confirmed the association of PI-RADS 5 with OS in men with a confirmed prostate cancer diagnosis. The Lille cohort represents a contemporary PI-RADS v2 era cohort, consisting of men fit for surgery, diagnosed with localised or locally advanced non-metastatic PCa, and treated with RP. As expected, these surgically treated men had better survival outcomes than our broader cohort, which included all men with non-metastatic PCa. In the Lille cohort, only two PCa-related deaths among 301 RP-treated patients despite a median follow-up of over six years were found. Nonetheless, PI-RADS remained a prognostic factor across both cohorts, particularly for MFS and BCR.

Our findings underscore that PI-RADS score 5 is a robust predictor of PCSM, additive to established prognostic markers such as biopsy GG and metastatic spread rather than merely serving as a surrogate marker for metastatic spread. Even after adjusting for these clinical confounders, the prognostic value of PI-RADS 5 remained statistically significant, highlighting its potential role as an independent imaging biomarker.

Interestingly, immediate curative-intent interventions (RP or RT) were not statistically significant predictors of PCSM in our analysis. This result aligns with findings from the ProtecT trial^35^, which reported no significant PCSM survival benefit between active monitoring and RP or RT. However, when considering OS, expected confounders such as age and immediate interventions became statistically significant. This likely reflects selection bias, where healthier patients are more likely to receive curative treatment.

Our findings may help identify a very high-risk group, such as GG 5 and PI-RADS 5 who may benefit from adjuvant or neoadjuvant therapies in addition to curative-intent treatment. Our data suggest that more intensive treatment may be warranted for men with PI-RADS 5. However, for men with otherwise favorable clinical parameters, active surveillance is sufficient. This is supported by recent findings by Peyrottes et al., who reported that although active surveillance remains feasible for PI-RADS 5 patients, these men have a higher likelihood of GG reclassification and earlier conversion to curative treatment.^36^ Our findings also have direct implications for PCa screening protocols. Men with PI-RADS 5 but negative biopsies should be considered for shorter re-screening intervals (1-2 years) to ensure that clinically significant cancers are not missed. Men with no suspicious findings, can likely be safely re-screened at longer intervals (4-6 years), thus optimizing the balance between early detection and overdiagnosis.

This study has several limitations that should be acknowledged. Its retrospective design and reliance on electronic health records introduced potential biases, particularly regarding metastatic status imaging data. This potentially influenced the analysis of metastatic spread in the HUS cohort in which 570 patients with PI-RADS scores of 3-5 had unknown biopsy status. This may reflect biopsy-related risks, particularly in elderly or comorbid patients, or indicate that biopsies were performed outside our healthcare system. Notably, unknown biopsy status was a significant predictor for OS, suggesting that these patients were more likely to die from other causes than PCa, and thus they would be unlikely to benefit from PCa treatment. Therefore, a prostate biopsy would not be considered necessary in such cases.

Despite the limitations, our study has several notable strengths. It includes three tertiary care centers with equal access to healthcare. It represents the longest follow-up study on PI-RADS v2 published to date. It includes detailed laboratory measurements, treatment data, comorbidities, and cause-specific mortality by linking data to national registries. The use of the Finnish National Cause-of-Death Registry ensured highly accurate PCSM endpoints.

Future research should focus on integrating AI-based tools into MRI interpretation and risk prediction ^37,38^. Deep learning models might enhance MRI-based prostate cancer diagnostics, which could lead to more standardized and objective assessments beyond PI-RADS scoring.

## Conclusions

We identified PI-RADS v2 score 5 as a robust predictor of PCSM, independent of biopsy GG and metastatic spread. PI-RADS 5 was also strongly associated with OS, MFS, and BCR.

Our findings were validated in both national (Tampere) and international (Lille) cohorts, highlighting the generalizability of PI-RADS as a prognostic imaging marker. The identification of a powerful MRI-based prognostic tool underscores the clinical importance of standardizing MRI scoring. These results reinforce the role of MRI in contemporary PCa risk stratification and management.

## Data Availability

All data produced in the present study are available upon reasonable request to the authors.

