## Supplementary Material for "PI-RADS v2 is a Strong Prognostic Marker for Adverse Outcomes in Prostate Cancer"

### Supplementary Figures

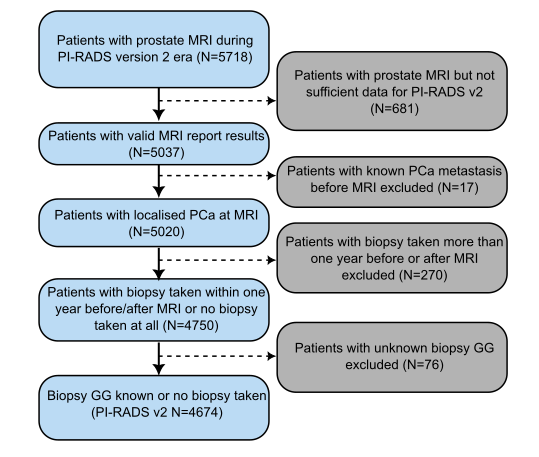

**Supplementary Figure S1**: Patient selection/exclusion at HUS.

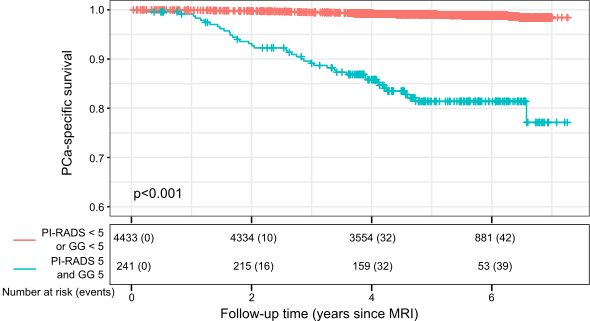

**Supplementary Figure S2**: Kaplan-Meier for patients with both PI-RADS 5 and GG 5 versus the rest in respect to PCa-specific survival. Reported p-values are log-rank tests.

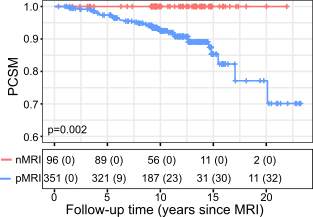

**Supplementary Figure S3**: Kaplan-Meier for pre-PI-RADS era patients and PCa-specific mortality in HUS.

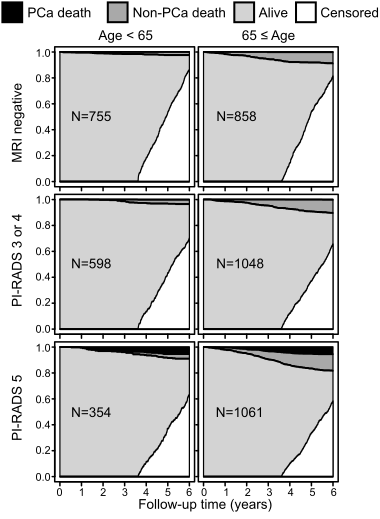

**Supplementary Figure S4**: Patient proportions stratified by age and PI-RADS during follow-up for competing risks with alive, censored, non-PCa death and PCa-specific in HUS.

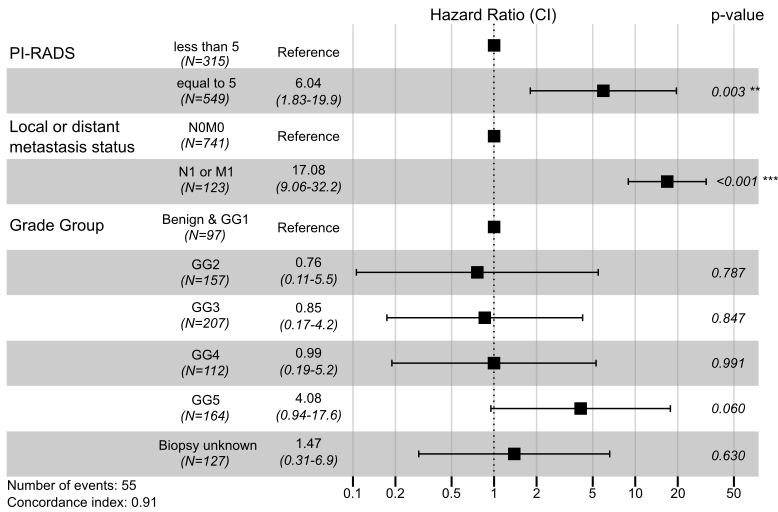

**Supplementary Figure S5**: Multivariable Cox model forest plot for PCa-specific mortality with PI-RADS, metastatic status, and biopsy Grade Group (GG) at MRI.

### Supplementary Tables

**Supplementary Table S1**: No biopsy or unknown versus PI-RADS version 2 score distribution, with absolute patient counts together with rounded total percentages in parentheses.

| **PI-RADS v2 scores for “no biopsy or unknown” reported patients *(N=1698)*** | | | | | |
| --- | --- | --- | --- | --- | --- |
| **0** | **1** | **2** | **3** | **4** | **5** |
| 1099 (65%) | 1 (0%) | 28 (2%) | 166 (10%) | 163 (10%) | 241 (14%) |

**Supplementary Table S2**: Competing risk estimates in HUS with the Fine-Gray model.

| **Event: Non-PCa death** | **HR** | **CI 95%** | **p-value** |
| --- | --- | --- | --- |
| Negative MRI | Reference | - | - |
| PI-RADS 3 or 4 | 1.55 | [1.15, 2.07] | 0.004 |
| PI-RADS 5 | 2.03 | [1.49, 2.79] | <0.001 |
| GG ≤ 1 | Reference | - | - |
| 2 ≤ GG ≤ 4 | 1.07 | [0,78, 1.47] | 0.660 |
| GG 5 | 1.40 | [0.90, 2.220] | 0.140 |
| GG Unknown | 1.60 | [1.20, 2.13] | 0.002 |
| **Event: PCa Death** | **HR** | **CI 95%** | **p-value** |
| Negative MRI | Reference | - | - |
| PI-RADS 3 or 4 | 4.93 | [0.56, 43.7] | 0.150 |
| PI-RADS 5 | 75.1 | [10.2, 554] | <0.001 |
| GG ≤ 1 | Reference | - | - |
| 2 ≤ GG ≤ 4 | 1.62 | [0.56, 4.70] | 0.370 |
| GG 5 | 10.1 | [3.63, 28.1] | <0.001 |
| GG Unknown | 3.50 | [1.19, 10.3] | 0.023 |

### Supplementary Methods

**HUS pre-PI-RADS era cohort**

For an additional validation with longer follow-up (median follow-up of 10.2 years) an additional pre-PI-RADS era cohort from HUS was identified. 447 men with MRI before the PI-RADS era (before 2012) were identified at HUS Acamedic and labelled as nMRI (no suspicion of clinically significant PCa, n=96) or pMRI (suspicion of clinically significant PCa in MRI, n=351).

**Pre-processing pathological findings for patient biopsies**

In the Helsinki University Hospital (HUS) cohort, we classified biopsies with no pathological findings as GG0. Moreover, we defined “No biopsy or unknown” patients as those whose biopsy data was not available in the pathology registry. In total, we identified 1,698 patients with an “No biopsy or unknown” biopsy status (Supplementary Table S1). Among these 1,698 patients in the PI-RADS v2 era, 1,128 patients (65%) had negative MRI results, 329 (19.4%) had PI-RADS scores of 3 of 4, and 241 (14.2%) patients had PI-RADS score 5 (Supplementary Table S1). The absence of biopsy results in patients with positive MRI findings is likely due to factors such as a high risk of infection, significant comorbidities, or advanced age, which may have precluded biopsy. Additionally, some biopsies may have been performed outside the HUS electronic health record (EHR) system.

**Collection and curation of patients’ metastasis status based on their bone and PET scans in the HUS cohort**

**Data collection**

- Queried all medical imaging acquisition numbers (AC numbers) and related reports in the HUS EHR system from 1999-2019 (data collected by L-Force Ltd).

**Curation process**

- Identified bone scan and PSMA/Choline-PET-CT visits and corresponding reports in the HUS EHR using the Nordic Classification of Surgical Procedures (NCSP) system:
  - NK6AN - bone scan (1999-2019)
  - KE1DR - PSMA-PET and NK6DR - Choline PET (2011-2019)
- Filtered all imaging referrals for men above 30 years old.
- Manually reviewed all referrals to ensure investigations were conducted for prostate cancer diagnosis or follow-up, using keywords such as spreading, metastasis, etc.
- All reports were signed out by nuclear medicine specialists with at least six years of experience at the time of reporting.

**Metastasis status determination**

- Bone scan: Patients were classified as metastasis-positive if at least one metastasis was mentioned in the report
- PSMA PET: Patients were classified as metastasis-positive if at least one metastasis was detected outside the prostate.
- Manual review: One author (Kerro O.) manually verified all reports for metastatic status.

**Pirkanmaa hospital district (Tampere University Hospital) data collection and curation**

A total of 17,000 patients diagnosed with prostate cancer (ICD-10 code C61) and treated in the Pirkanmaa Hospital District were retrospectively identified from the EHR system of Tampere University Hospital.

Among these patients, 1,587 had PI-RADS scores recorded in the free-text reports. However, only 1,564 of these patients had a corresponding MRI imaging record.

Date of birth and, if applicable, date of death was extracted from structural reports. Birth dates were available for 1,484 patients. Follow-up period started on the date of the MRI. The follow-up period ended on the date of death or at the end of the study period (December 31, 2021), whichever occurred first. 10 patients with a follow-up time of 30 days or less were excluded, resulting in a final cohort of 1,474 patients.

PSA measurements were available for 1,371 patients. PSA values were extracted from structured data and free-text pathology reports. Only PSA values recorded within 90 days of MRI imaging were included.

Biopsy GG was available for 1,351 patients. GG was extracted from structured and free-text pathology reports. Only biopsies performed within one year of MRI imaging were included. Both PSA values and biopsy GG were collected before curative treatment (RP or RT). Key demographics of the Tampere University Hospital cohort are presented in Table 1.

**Lille University Hospital data collection and curation**

A total of 1,864 treatment-naïve patients with nonmetastatic PCa were collected from Lille University Hospital between 2009 and 2018. All patients underwent pre-biopsy MRI, a 12-core transrectal ultrasound guided systematic biopsy, and two targeted biopsies, and provided informed consent for the use of their personal data for research purposes.

All patients were prospectively followed, with detailed clinical data collected throughout the study period. Among these patients, 713 patients underwent RP within one year after diagnosis, had a recorded MRI, a follow-up time of over 18 months, and did not receive adjuvant radiation therapy. To ensure inclusion of only patients with MRI recorded under the PI-RADS v2 reporting system, patients were selected if their MRI was performed on or after January 1, 2015.

After applying these criteria, 301 patients were included in the study, with median age of 68 years (IQR 64-84) and median follow-up of 6.2 years (IQR 4.8-7.4).

PSA values and age were recorded at the time of diagnosis. BCR and metastasis were closely monitored throughout the follow-up period. BCR was defined as a PSA rise >0.2 ng/mL. Metastasis-positive status was recorded if at least one suspicious node (>1 cm) or bone lesion was detected on MRI or PET/CT scans. For further details on the dataset, refer to Bommelaere et al.^1^ Key demographics of the Lille University Hospital cohort are presented in Table 1.
